# Validation of diagnostic nomograms based on CE-MS urinary biomarkers to distinguish clinically significant prostate cancer

**DOI:** 10.1101/2022.01.26.22269739

**Authors:** Maria Frantzi, Isabel Heidegger, Marie C. Roesch, Enrique Gomez-Gomez, Eberhard Steiner, Antonia Vlahou, William Mullen, Ipek Guler, Axel S. Merseburger, Harald Mischak, Zoran Culig

## Abstract

**Background:** Prostate cancer (PCa) is the most common cancer and one of the leading causes of death worldwide. However, a significant proportion of PCa are low risk PCa which do not require an active treatment due to its low mortality rates. Thus, one major issue in PCa management is to accurately distinguish between indolent and clinically significant (cs) PCa to reduce overdiagnosis and overtreatment. In this study, we aim to validate the performance of diagnostic nomograms (DN) based on previously published urinary biomarkers for discriminating csPCa.

**Patients and Methods:** Capillary electrophoresis/ mass spectrometry has been employed to validate a published biomarker model based on 19 urinary peptides specific for csPCa. Added value of the 19-biomarker model (19-BM) was assessed in diagnostic nomograms including prostate specific antigen (PSA), PSA density and the risk calculator from The European Randomized Study of Screening for Prostate Cancer (ERSPC). For this purpose, urine samples from 147 PCa patients (including 80 low, 44 intermediate and 17 high risk patients) were collected prior to prostate biopsy. The 19-BM score was calculated via a support vector machine-based software based on the pre-defined cut-off criterion of -0.07. DNs were subsequently developed to assess added value of integrative diagnostics.

**Results:** Independent validation of the 19-BM resulted in 87% sensitivity and 65% specificity, with an AUC of 0.81, outperforming PSA (AUC_PSA_:0.64), PSA density (AUC_PSAD_: 0.64) and ERSPC-3/4 risk calculator (0.67). Integration of 19-BM with the other clinical variables into distinct DN, resulted in improved (AUC range: 0.82-0.88) but not significantly better performance compared to 19-BM alone.

**Conclusions:** 19-BM alone or combined with clinical variables into DN, demonstrated value for detecting csPCa, and decreasing the number of biopsies.

## Introduction

Prostate Cancer (PCa) ranks as the second most frequent and the fifth leading cause of cancer death among men. In 2020, almost 1.4 million new cases were diagnosed worldwide, and 375,000 deaths were reported due to PCa [1]. Although this malignancy is diagnosed in 15-20% of men, the lifetime risk of death is significantly lower (3%) [2]. PCa patients represent a heterogeneous group, with many of them presenting slow growing forms of PCa, unlikely to progress in the absence of treatment and others having aggressive life-threatening disease if left untreated. For patients presenting with slow growing PCa, defined also as clinically insignificant cancer (insPCa: Gleason score-GS < 7 and PSA < 10 ng/ml) [3], immediate treatment is not recommended, rather a conservative management by active surveillance (AS) [3].

Screening for PCa is currently based on serum prostate specific antigen (PSA) testing and digital rectal examination (DRE). However, multiple factors not related to prostate malignancy may affect the level of blood PSA (inflammation, infection, presence of benign prostate hyperplasia etc.) [4]. Therefore, PSA lacks specificity, with only ∼40% of all patients with an elevated PSA positively confirmed with PCa following biopsy [5]. The introduction of intensive PSA screening led on one hand to early detection of PCa, on the other hand numerous insPCa are diagnosed often associated with overtreatment. According to the European Association of Urology (EAU) guidelines [3], definitive diagnosis of PCa is based on the histopathological confirmation of PCa in biopsy cores, following a positive result of DRE and/ or high PSA levels [3]. Until recently, the procedure was guided by transrectal ultrasound (TRUS) upon administration of local anesthesia [6]. TRUS guided biopsy is an invasive procedure associated with several side effects like infectious complications, hematuria, bleeding episodes and urinary clot retention [7]. In an effort to improve the accuracy for PCa detection, multiparametric magnetic resonance imaging (mpMRI) has been recently adopted, resulting in good sensitivity for detecting GS ≥ 3+4 (sensitivity of 91%, specificity of 37%) [8], although it is less sensitive for GS < 3+4 (sensitivity of 70%, specificity of 27%) [8]. While mpMRI is beneficial, particularly for guiding repeated biopsy [9], inter-reader variability among radiologists as well as the limited capacity to perform a high number of MRI-guided procedures remain significant challenges [10].

Considering the above challenges, a non-invasive test to guide not only initial but also repeated biopsies would be of added value. High throughput *-omics* technologies have enabled simultaneous analysis of thousands of features and a better definition of molecular pathophysiology in cancer [11]. In the context of PCa, although several candidate biomarkers have been described [12], single biomarkers frequently lack diagnostic accuracy for routine clinical application due to disease heterogeneity [13]. The high biological variability of PCa suggests that a combination of multiple, *-omics* derived biomarkers into integrative diagnostics, rather than a single biomarker, are better suited to accurately detect significant PCa. In this context, high resolution urinary proteomics profiles from >800 patients have been acquired by capillary electrophoresis coupled to mass spectrometry (CE-MS). Subsequently, proteomics patterns that were developed using machine learning algorithms in a form of a 19-biomarker model (19-BM), have been used to discriminate csPCa (GS ≥7) from slow-progressing PCa in patients with low PSA levels (<15 ng/mL) [14]. Based on the previously published data [14], the 19-BM resulted in a 90% sensitivity and 59% specificity, with an AUC of 0.81, outperforming PSA (AUC_PSA_: 0.58) and the ERSPC-3/4 risk calculator (AUC_ERSPC_: 0.69). Moreover, based on a first investigation, integration of the CE-MS biomarkers with other variables like PSA and age showed an increased performance (AUC: 0.83), demonstrating a level of complementarity of these variables. Considering this evidence, in this study, the aim was to validate the previously established CE-MS-based 19-BM, and additionally investigate whether integrative models, including 19-BM in combination with current state-of-the art clinical risk calculators can lead to improved non-invasive discrimination between insPCa and csPCa.

## Methods

### Patient population and characteristics

This study was performed according to the REMARK Reporting Recommendations [15] and the recommendations for biomarker identification and reporting in clinical proteomics [16], including 148 patients who underwent a transrectal ultrasound (TRUS) -guided prostate biopsy according to clinical guidelines at the Department of Urology in Innsbruck Medical University, between 2016 and 2018, as part of BioGuidePCa project (E! 11023, Eurostars). Sample collection and processing were ethically approved by the local ethics committee at Innsbruck Medical University and informed consent was obtained from all participants. Of the 148 patients for whom biopsy results confirmed presence of adenocarcinoma of prostate, one patient was excluded as the PSA measurement was missing. A schematic representation of the study design is presented in **Figure 1**. D’Amico classification used Gleason Score (GS) and PSA criteria as per D’Amico *et al*. [3, 17]. At the time of patient enrollment, mpMRI-guided biopsy was not yet implemented in the clinical practice, therefore all patients underwent TRUS-guided biopsy to provide biopsy information. TRUS-guided prostate biopsy was carried out under local anesthesia by using a standard periprostatic block, a TRUS transducer, and an 18-gauge automated needle biopsy instrument. The prostatic volume was measured, and 10 biopsy cores were obtained. Full clinical and laboratory data, including among others: PSA level, the results of DRE, prostatic volume, number of previous biopsies and GS were collected and are presented in the **Supplementary Table S1**. The European Randomised Study of Screening for Prostate Cancer (ERSPC) estimates for risk stratification were calculated as previously described [18], considering serum PSA levels, the DRE result and information about the previous biopsies. The patient cohort characteristics are summarized in **Table 1**.

**Table 1.** Clinical and biochemical variables for the 147 patients with confirmed PCa

| **Baseline characteristics** | **Patients with confirmed PCa (n=147)** |
| --- | --- |
| **Median age** (95% CI; yr) | **66.0** (64.2-67.0) |
| **Age range** (yr) | **40 - 84** |
| **PSA median** (95% CI; ng/ml) | **5.2** (4.5-5.9) |
| **Digital Rectal Examination**<br>(normal/ suspicious / NA) | **90 /20 / 37** |
| **Previous biopsies** (Y/N) | **43/ 104** |
| **Median urinary creatinine**<br>(95% CI; mmol/L) | **8.8** (7.6 - 10.3) |
| **Median total protein**<br>(95% CI; mmol/L) | **0.03** (0.03 - 0.05) |
| **Disease pathology** |  |
| ▪ GS 6 | **99** |
| ▪ GS 3+4 / 4+3 | **31/ 4** |
| ▪ GS 8 | **8** |
| ▪ GS 9 | **5** |
| **D'amico risk Stratification** |  |
| ▪ Low risk | **80** |
| ▪ Intermediate risk | **44** |
| ▪ High risk | **17** |
| **Significant PCa (GS ≥ 3+4)** | **48** (32.7%) |
| **Insignificant PCa (GS: 6)** | **99** (67.3%) |

**Abbreviations:** 95% CI: Confidence interval; GS: Gleason Score; NA: data not available; N: Not received; PCa: Prostate Cancer; Y: Received

**Figure 1:**
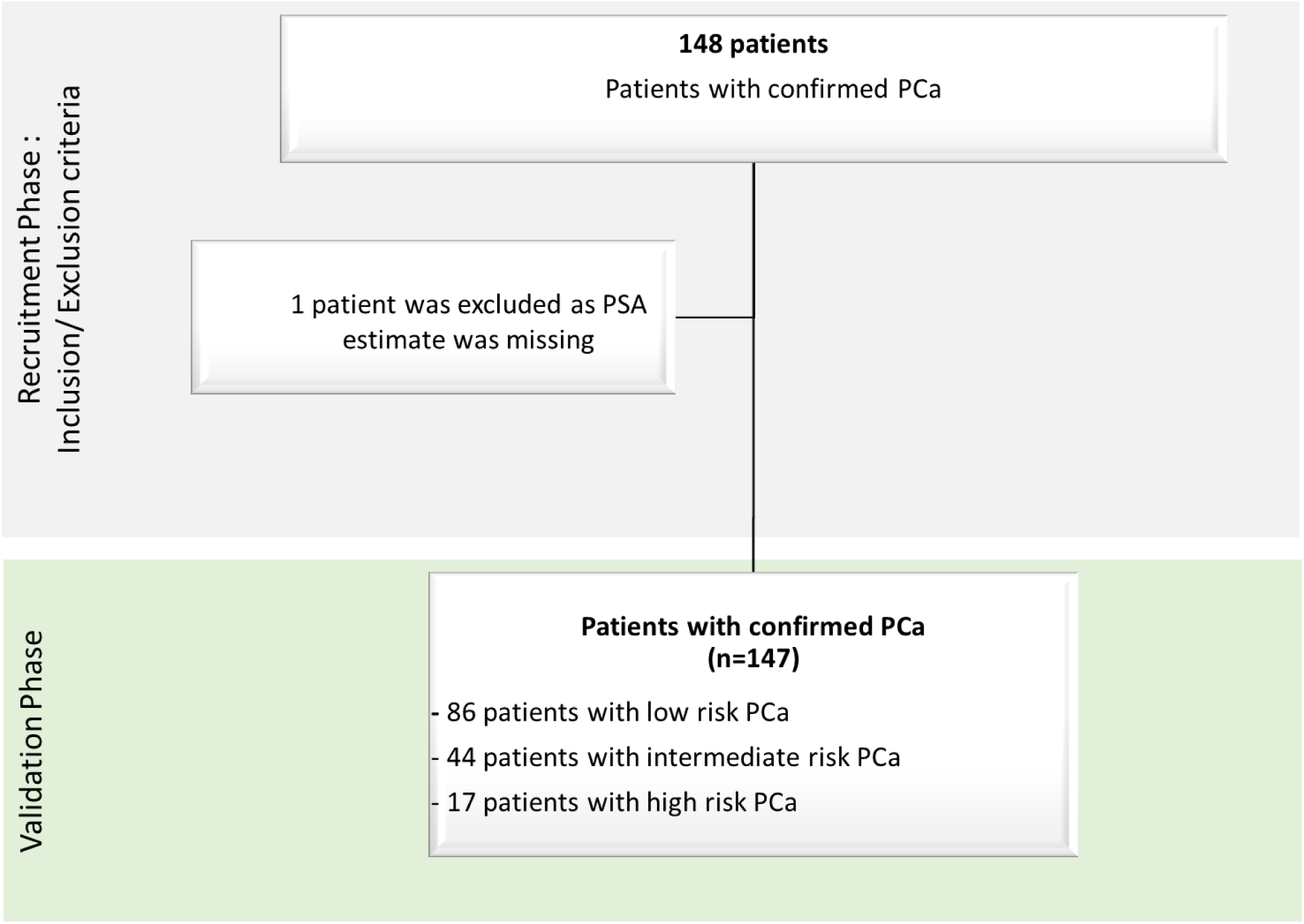
Schematic representation of the study design for the validation of urine CE-MS based nomograms.

### Urine collection and processing

All urine samples were collected prior to prostate biopsy according to clinical guidelines. Voided urine samples were collected in sterile containers and immediately stored at -20°C until further processing. Sample preparation was performed by diluting 700 µl aliquots from the urine collected from PCa patients after DRE, in two volumes (1:2) alkaline buffer containing 2M urea, 10mM NH_4_OH and 0.02% SDS (pH 10.5). The samples were subsequently filtered by Centrisart ultracentrifugation filters (Sartorius, Göttingen, Germany) to retain proteins/ polypeptides below 20kDa that were further desalted through PD-10 columns (GE Healthcare, Munich, Germany).

### Mass Spectrometry analysis

CE-MS analysis was performed in 147 urine samples, following the previously established protocols for samples preparation and data acquisition [19]. CE-MS analysis and data processing was performed according to ISO13485 standards yielding quality controlled urinary data sets [20]. Mass spectral ion peaks representing identical molecules at different charge states were de-convoluted into single masses using MosaiquesVisu software [20, 21]. A peak list of each peptide was defined by its molecular mass [kDa], normalized migration time [min] and normalized signal intensity [AU] [21]. Normalization of the CE-MS data was based on 29 internal collagen fragments stable over disease/ health state that serve as internal standards [22]. All detected peptides were deposited, matched, and annotated in a Microsoft SQL database [23] and used as input in the presented study. These data have not been previously described before and are unique to this study. Transformation of the data (log-transformation) was performed before performing the statistical analysis, as previously described [24].

### Statistical evaluation of model predictivity

The proportion, mean, standard deviation, median and interquartile range (25th–75th percentiles) estimates were calculated to describe the distribution of the different variables in the patient cohort (summarized in **Table 1**). The biomarker model’s scores were calculated via the support vector machine (SVM)-based software, namely MosaCluster (version 1.7.0), as previously described [25]. The sensitivity and specificity estimates for the SVM-based peptide marker pattern were calculated based on the number of correctly classified samples, as defined by biopsy, considering the previously reported cut-off criterion of (−0.07). The receiver operating characteristic (ROC) plots and the respective confidence intervals (95% CI) were based on exact binomial calculations and were calculated in MedCalc 12.7.5.0 (Mariakerke, Belgium). The area under the ROC curve (AUC) was evaluated to estimate the overall accuracy independent upon a particular threshold [26], and the values were then compared using DeLong tests. Statistical comparisons of the classification scores between the PCa risk groups and GS groups were performed by the Kruskal-Wallis rank sum test using MedCalc 12.7.5.0 (Mariakerke, Belgium) [27]. The diagnostic nomograms (DN) of 19-BM in combination with clinical variables were established using multiple linear regression analyses. Decision curve analysis (DCA) [28] examined the potential net benefit of using the diagnostic nomograms in the clinic, according to which a net benefit is defined as a function of the decision threshold at which one would consider obtaining a biopsy. The list of scoring data is presented in **Supplementary Table S2**.

## Results

### Patient characteristics

In this study, CE-MS proteomics analysis was performed in urine samples from 148 patients, for which biopsy results confirmed presence of prostate adenocarcinoma. Of these 148, one patient had to be excluded as PSA values were not available. The PCa patients were classified into risk groups according to D’Amico criteria [17] and EAU guidelines [3], revealing 86 patients of low, 44 of intermediate and 17 of high risk (**Table 1**). Patients in the low risk group were of median age of 64 years (61.9-66.0; 95% CI), while the median value of PSA was 4.4 (4.1-4.7; 95% CI). Accordingly, PCa patients of intermediate risk presented median age of 69 (65.6-71.4; 95% CI), while the median value of PSA was 6.9 (5.5-9.4; 95% CI). Finally, seventeen high risk PCa patients were classified with median age of 72 (67.0-74.0; 95% CI) and median PSA levels of 18.1 ng/ml (9.7-20.9; 95% CI). Out of 147 patients, 99 (67.3%) presented with insignificant (GS: 6) and 48 (32.7%) with significant PCa (GS ≥ 3+4). Men with csPCa were significantly older (median age of 69.5; 67.0-72.0; 95% CI) compared to men with insPCa (median age of 65; 62.4-66.0; 95% CI; *p= 0.0053; Mann-Whitney test*) and had significantly higher PSA levels [(PSA_ins._= 4.6 (4.2-5.3; 95% CI) compared to PSA_sig._=6.6 (5.2-8.8; 95% CI); *p=0.0055; Mann-Whitney test*]. Moreover, out of 99 patients with insPCa, 70 (70,7%) did not undergo any previous biopsy, while 29 (29,3%) were previously biopsied. For those patients with csPCa (n=48), the respective proportion was 70.8% (n=34) who did not have previous biopsies and 29.2% (n=14) that had previously undergone biopsies. The difference in the percentage proportion of the number of previous biopsies between clinically cs and insPCa was not significant (*p=0.8565; Chi-squared test*).

### Validation of 19-BM based on CE-MS urinary peptide biomarkers

First the performance of the previously published 19-BM, based on high resolution CE-MS urinary profiles [14] was validated, in line with the recommendations for biomarker identification and reporting in clinical proteomics [16]. Considering 99 patients with ins.PCa and 48 patients with csPCa, the AUC for the 19-BM (AUC_19-BM_) was estimated at 0.803 with the 95% CI ranging from 0.73 to 0.86 (*p< 0.0001*). At the validated cut-off level of - 0.07 [14], the sensitivity was estimated at 87.5% and the specificity at 64.6% (**Figure 2A**). The 19-BM correctly classified 42 out of the 48 csPCa (GS ≥ 3+4) whereas 35 out of the 99 insPCa (GS: 6) were misclassified as csPCa cases. Considering a prevalence rate of 32.7 for csPCa, as estimated on the basis of this patient cohort, the negative predictive value (NPV) for detecting csPCa was computed at 91.8% (71.9-99.1%; 95% CI) while the positive predictive value (PPV) at 55.0% (34-75.8%; 95% CI). Moreover, as presented in **Figure 2B**, the 19-BM significantly discriminated between significant and insignificant PCa (*p <0.0001*, Kruskal-Wallis H test), separated patients with GS 6 from both those with GS 7 and GS ≥8 (*p < 0.005*, Kruskal-Wallis H test; **Figure 2C**) but also according to the risk group (low risk from intermediate and high risk; *p<0.005*, Kruskal-Wallis H test; **Figure 2D**). Additionally, the classification based on the urinary 19-BM was not affected by biochemical variables like urinary creatinine (Spearman’s rank correlation coefficient: 0.104; -0.06 to 0.27; 95% CI; *p=0.2278*) and total urinary protein (Spearman’s rank correlation coefficient: -0.058; -0.22 to 0.11; 95% CI; *p=0.5060*).

**Figure 2:**
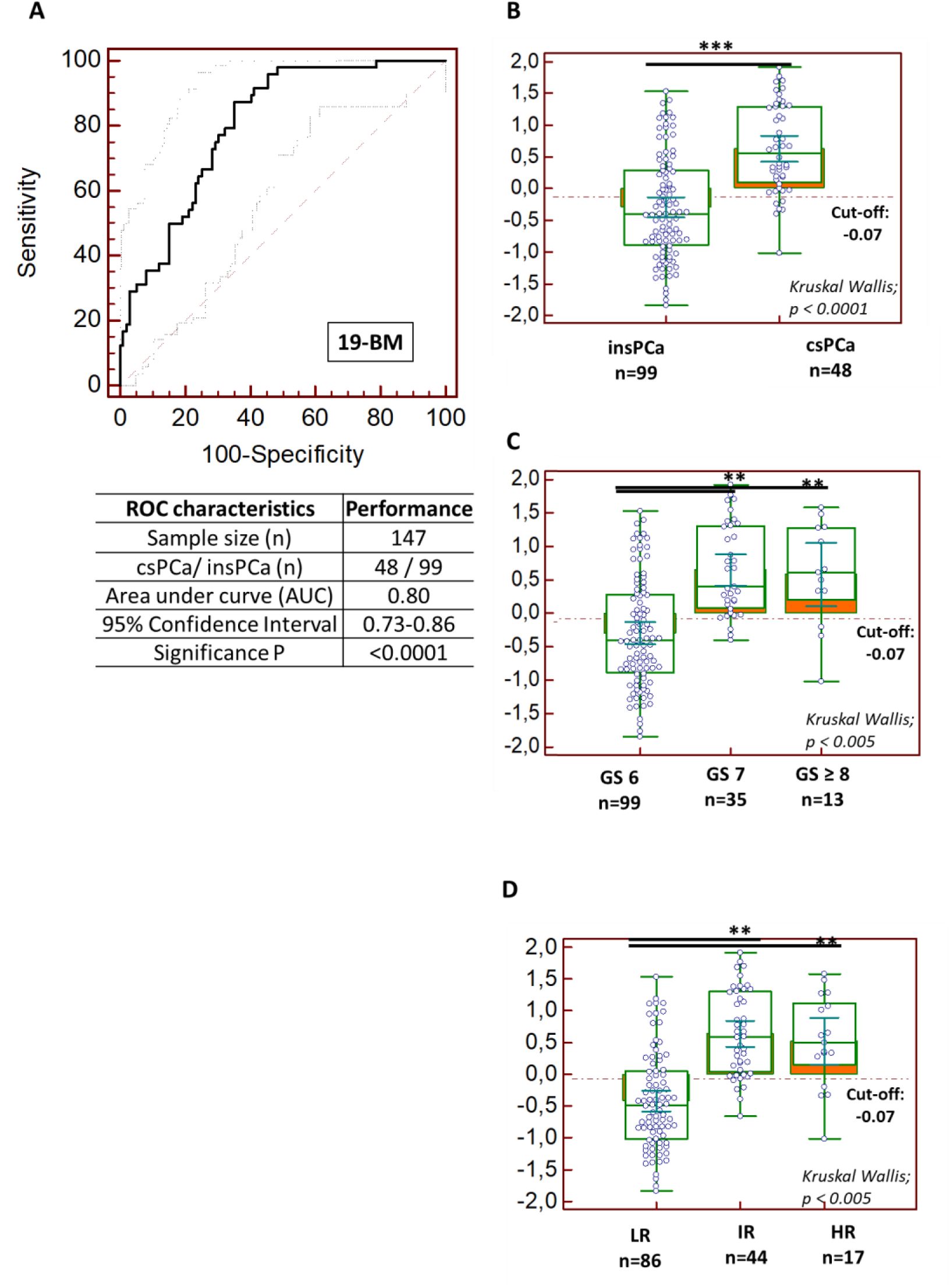
**A)** Receiver operating characteristics (ROC) analysis displaying the performance of the 19-biomarker model for discriminating csPCa from nsPCa; **B)** Classification scores, presented in Box-and-Whisker plots grouped according to the csPCa (n=48) and nsPCa (n=99), **C)** Classification scores displaying the level of discrimination across the different Gleason score, and **D**) risk groups based on D’amico classification. A post-hoc rank-test was performed using Kruskal-Wallis test.

### Comparative assessment of 19-BM with PSA and PSA density

Considering serum PSA levels that were measured at the timepoint of biopsy, a direct comparison of PSA and PSA density with the CE-MS based 19-BM was performed. Multivariate analysis showed that the 19-BM outperformed PSA in discriminating csPCa (AUC_PSA:_ 0.64;0.55-0.72; 95% CI; *p= 0.006;* **Figure 3A**). The sensitivity of the PSA was estimated at 66.7% (51.6-79.6%; 95% CI; PSA > 4ng/ml), while the specificity at 44.4% (34.5-79.6; 95% CI; PSA > 4ng/ml). Sixteen patients with csPCa were detected by the 19-BM but were missed by serum PSA. Out of 16 patients, 13 were bearing GS 3+4 PCa tumours, and three patients had tumors of GS ≥ 4+3. Additionally, both the NPV (73.3; 48.4-90.6; 95% CI) and the PPV (36.8; 19.5-57.0; 95% CI) estimates were lower than the estimates based on the 19-BM. Considering 135 PCa patients, for whom data on prostate volume were available, a direct comparison of the 19-BM with PSA density (PSAD) was also possible. As in the case of serum PSA, the 19-BM outperformed PSAD in discriminating csPCa (AUC_PSAD:_ 0.64;0.56-0.73; 95% CI; *p= 0.0127;* **Figure 3B**). The sensitivity of the PSAD was estimated at 83.7% (69.3-93.2%; 95% CI), with the specificity at 42.4% (32.1-53.1; 95% CI). Six patients with csPCa were detected by the 19-BM but were missed when considering PSAD. Out of these six patients, four were bearing GS 3+4 PCa tumours and two patients had tumors with GS ≥ 4+3. Similar to the PSA comparison, both the NPV (84.4; 55.6-97.8; 95% CI) and the PPV (41.3; 23.2-61.4; 95% CI) estimates for PSAD were lower than the estimates based on the 19-BM.

**Figure 3:**
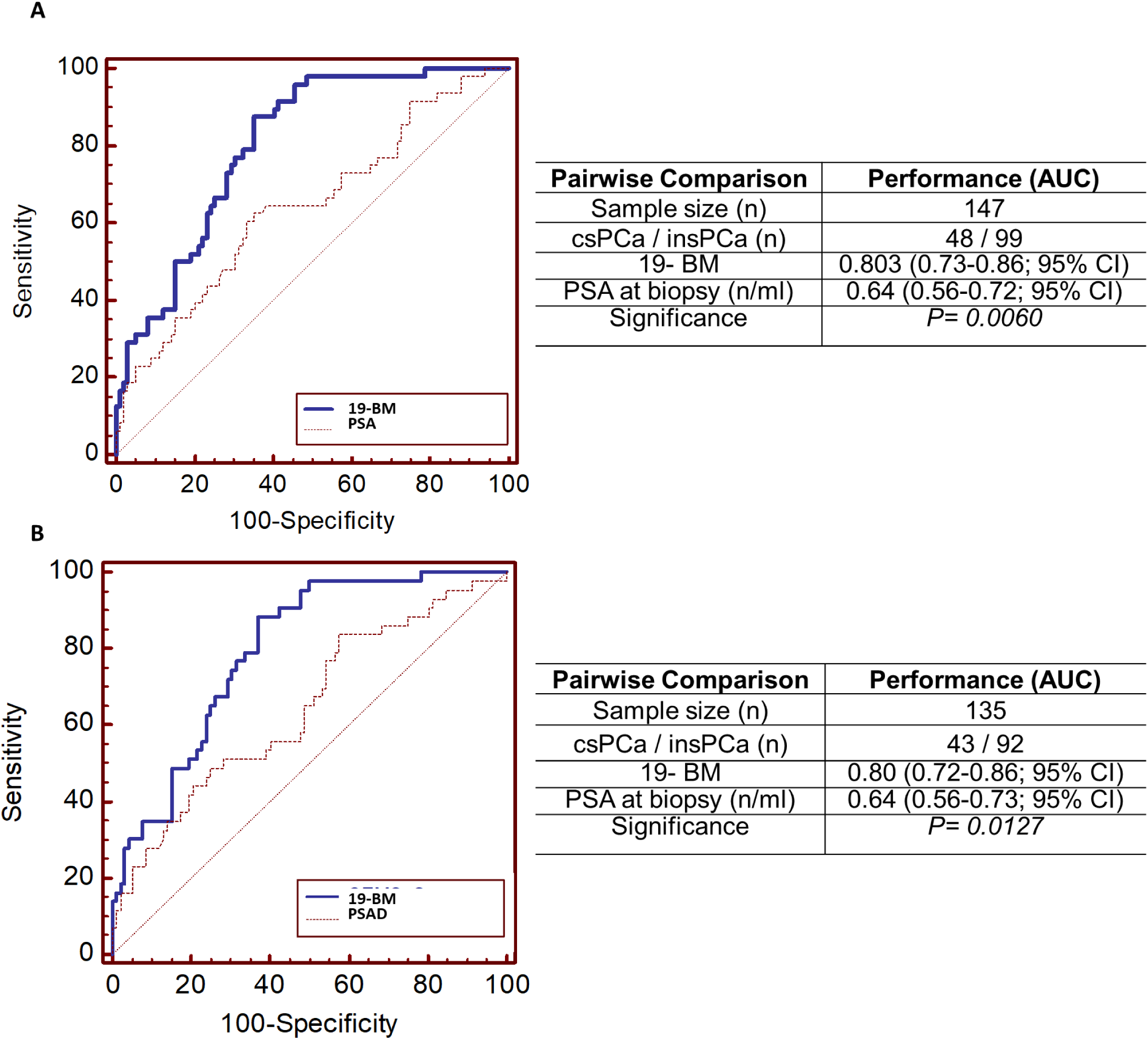
Comparative analysis depicted by receiver operating characteristics (ROC) curves of the 19-biomarker model (19-BM) with **A)** serum PSA measurements and **B)** PSA density (PSAD)

### Comparison of 19-BM with the ERSPC clinical risk calculator

To investigate if the 19-BM can improve on the current state-of-the-art clinical prognosticators, the SVM-based scores from 19-BM were further compared with the estimates of ERSPC risk calculator for detecting high risk PCa (ERSPC 3/4), as presented in **Figure 4**. Based on the available clinical data for the PSA levels, the DRE result and accounting also for the previous biopsies, ERSPC 3/4 estimates for 109 PCa patients were available for this comparison. In this analysis the performance of 19-BM (_AUC19-BM_=0.82; 0.74-0.89) was significantly superior to the one of ERSPC 3/4 risk calculator (AUC_ERSPC3/4_ =0.67; 0.57-0.76; *p= 0.0275*). At the optimal cut-off level sensitivity of the ERSPC 3/4 was estimated at 52.9% (35.1-70.2; 95% CI) and specificity at 72.0% (60.4-81.8; 95% CI). Fifteen of the 34 patients with confirmed csPCa, including three GS≥8 tumours were misclassified by the ERSPC3/4. Interestingly, fourteen of the 15 were detected by the 19-BM. Considering the predictive values, both the NPV (75.9; 52.5-91.6; 95% CI) and the PPV (47.9; 19.6-77.3; 95% CI) estimates based on ERSPC 3/4 were slightly but not significantly lower than the estimates based on the 19-BM.

**Figure 4:**
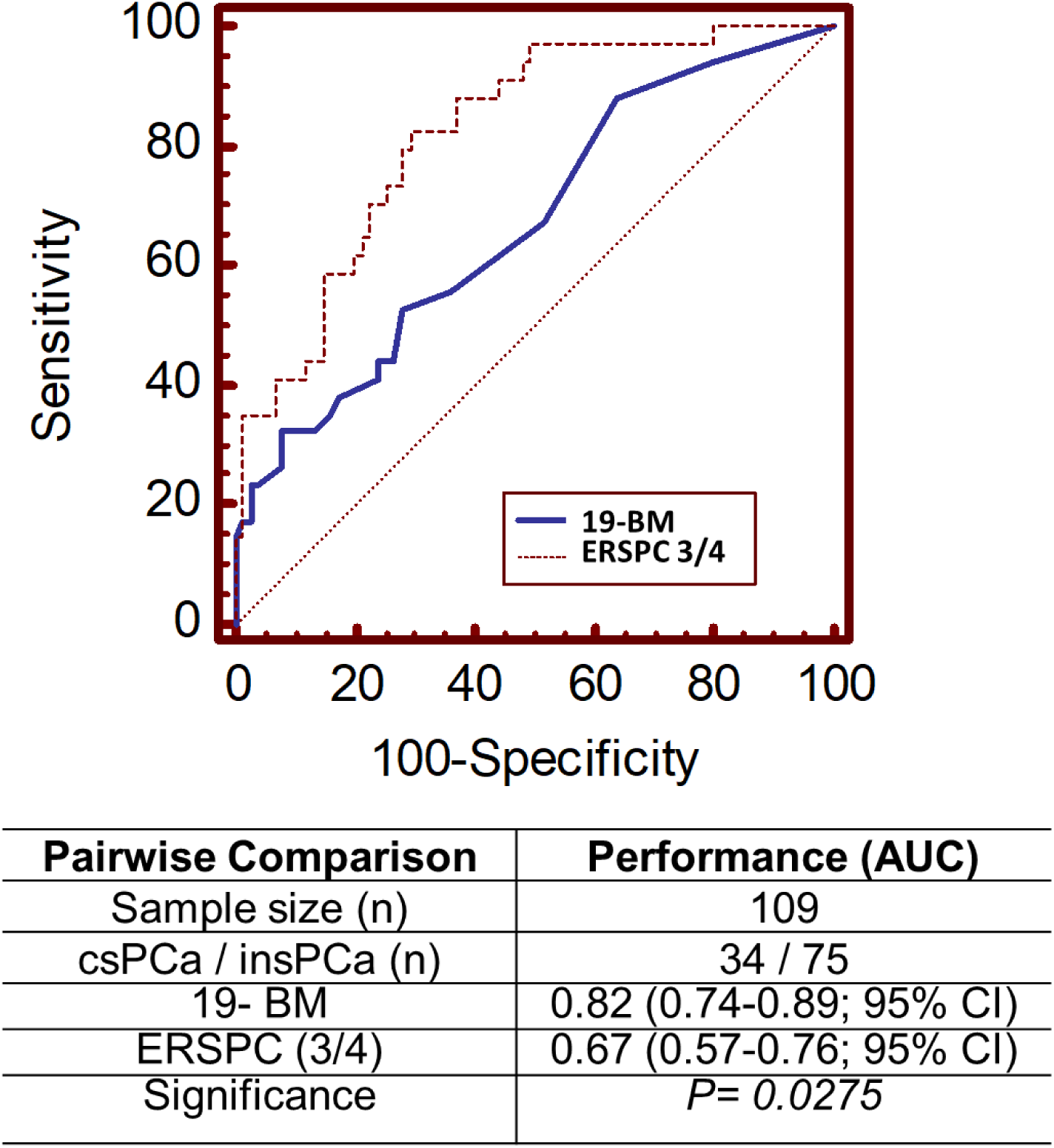
Comparative analysis depicted by receiver operating characteristics (ROC) curves for the 19-BM and the ERSPC 3/ 4, considering a subgroup of 109 PCa patients for whom, available clinical data enabled ERSPC estimation (34 csPCa and 75 insPCa).

### Integrative diagnostics: Assessment of multimodal models based on 19-BM

Evidence based on initial analysis of previously reported data [14] showed a slightly improved performance upon integration of 19-BM to a DN including CE-MS based 19-BM, PSA and age (DN_PSA-AGE-19BM_). As before [14], in this study integration of PSA and age with the 19-BM, DN_PSA-AGE-CE_, resulted in an improved AUC value of 0.83 (0.76-0.89; 95% CI), although not statistically significant (*p= 0.1308*) compared to the 19-BM (AUC: 0.80; 0.73-0.87; **Figure 5A**). The integrative nomogram outperformed PSA (AUC: 0.64; 0.5-0.72; 95% CI; *p= 0.0001*; **Table 2**). A second DN was developed by integrating PSAD and 19-BM (DN_PSAD-19BM_), performing again slightly (AUC_DN:PSAD-CE_: 0.82; 0.74-0.88; 95% CI) but not significantly better than 19-BM alone (*p= 0.2421;* **Figure 5A**; **Table 2**). In comparison to PSAD alone, DN_PSAD-19BM_ demonstrated significantly better performance based on the ROC pairwise comparison (*p= 0.001)*. Similarly, a third DN was evaluated by combining ERSPC 3/4 with 19-BM (DN_ERSPC3/4-19BM_). In this assessment, too, and although the integrative performance for DN_ERSPC3/4-19BM_ was even higher reaching an AUC of 0.86 (0.78 – 0.92; 95% CI), the difference was not statistically significant when compared to 19-BM alone (*p= 0.076;* **Figure 5A**; **Table 2**). Lastly, all the above significant clinical variables were integrated together into a DN including all relevant risk estimates such as the 19-BM, PSA, PSAD, age and ERSPC 3/4. As shown in Figure 5A, the performance for this DN including all significant variables was further improved reaching an AUC of 0.88 (0.80-0.93; 95% CI), significantly outperforming PSA (*p= 0.002*), PSAD (*p < 0.001*), ERSPC 3/4 (*p= 0.0007)* alone but not the 19-BM (*p= 0.06*) alone. Considering the optimal cut-off criterion for the diagnostic nomogram (>0.1766; Youden index), the sensitivity was estimated at 93.8% (79.2 – 99.2; 95% CI) and the specificity at 65.7% (53.4-76.7; 95% CI). The integrative nomogram correctly classified 30 out of the 32 significant PCa (GS ≥ 3+4) and misclassified two patients with GS 7. Additionally, the NPV for detecting csPCa was computed at 95.6% (71.3-99.9%; 95% CI) whereas the PPV at 57% (32-79.6%; 95% CI). To assess the clinical benefit of the integrative model, a decision curve analysis was performed. Based on the net plotting against the threshold probabilities for the comparisons between the DN and the 19-BM, PSA, PSAD and ERSPC estimates, there is a clear benefit of the integrative model particularly in the lower range of the risk thresholds (**Figure 5B**).

**Table 2.** Summary of pairwise statistical comparisons for the developed integrative diagnostic nomograms (DN)

|  | **AUC** | **95% CI** | **P value** |
| --- | --- | --- | --- |
| **DN: PSA-age-19BM** | 0.83 | 0.76 to 0.89 |  |
| • 19BM | 0.81 | 0.73 to 0.87 | *0.1308* |
| • PSA | 0.64 | 0.56 to 0.72 | ***0.0001*** |
| **DN: PSAD-19BM** | 0.82 | 0.74 to 0.88 |  |
| • 19BM | 0.80 | 0.72 to 0.86 | *0.2421* |
| • PSAD | 0.64 | 0.56 to 0.73 | ***0.0001*** |
| **DN: ERSPC3/4-19BM** | 0.86 | 0.78 to 0.92 |  |
| • 19BM | 0.82 | 0.74 to 0.89 | *0.076* |
| • ERSPC 3/4 | 0.67 | 0.57 to 0.76 | ***0.001*** |
| **DN: all variables** | 0.88 | 0.80 to 0.93 |  |
| • 19BM | 0.82 | 0.73 to 0.89 | *0.0645* |
| • PSA | 0.69 | 0.60 to 0.78 | ***0.0002*** |
| • PSAD | 0.65 | 0.56 to 0.75 | ***< 0.0001*** |
| • ERSPC 3/4 | 0.69 | 0.59 to 0.78 | ***0.0007*** |

**Abbreviations:** 95% CI: Confidence interval; AUC: Area under the ROC curve; BM: biomarker model; DN: diagnostic nomogram; PSA: prostate specific antigen; PSAD: prostate specific antigen density; ROC – receiver operating characteristics.

**Figure 5:**
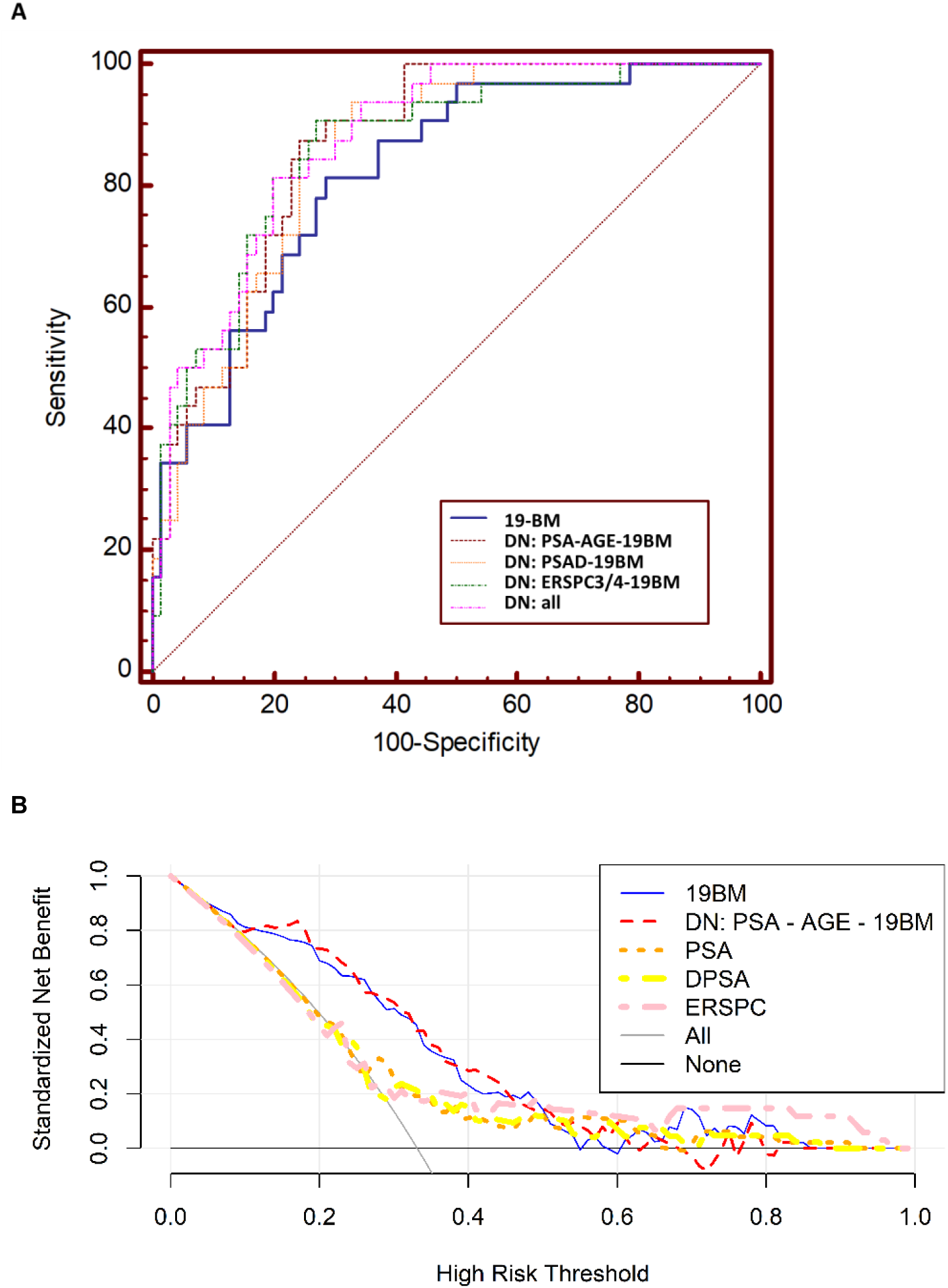
**A)** Receiver operating characteristics (ROC) analysis displaying the performance of DN nomograms based on 19-BM combined with the state-of-the-art risk variables (PSA, PSAD, ERSPC 3 /4 and age). **B)** Results of the decision curve analysis, comparing the net benefit for the prediction of csPCa on biopsy using the 19BM (blue line), and DN (PSA-AGE-19BM; red line), PSA (orange line), DPSA (yellow line) and ERSPC high risk 3/ 4 (pink line) as a function of the risk threshold, compared to those benefits of the strategies of treating all patients (grey line) and treating none (black line).

## Discussion

There is evidence that AS is a feasible and safe option for patients with low risk PCa. However, accurate and frequent monitoring is required to detect disease progressing to detect significant cancer in a timely fashion. Routine monitoring is commonly based on either PSA levels over time in addition to regular mpMRI and re-biopsy, the latter requiring the use of invasive needle biopsies [29]. To delay or even avoid regular re-biopsies for monitoring the disease, biomarkers predicting disease progression are of utmost importance. *-omics* based studies have been published reporting on predictive features of PCa biopsy outcome to guide patient stratification to significant and non-significant prostate cancer and improve patient management [30, 31]. Using CE-MS proteomics, a biomarker model based on 19 urinary peptides (19BM) was established and validated in 823 patients suspicious for presence of PCa (reporting an AUC of 0.81, outperforming PSA and ERPSC) [14]. Building upon this previous report, in this study we aimed at validating the 19-BM in 147 patients with confirmed PCa. The 19-BM demonstrated high reproducibility in this external validation study for discriminating significant PCa. The 19-BM exhibited good performance (AUC_19-BM_:0.80) that was comparable to the previously described estimates (AUC:0.81). Furthermore, the 19-BM sensitivity (88%) and specificity (65%) were similar to those previously reported (90% and 59%, respectively) [14]. The lower specificity is mostly attributed to the mis-classification of clinically insignificant PCa as clinically significant forms. The clinical consequence of this observation can be weighted as tolerable, since patients with positive score based on the CE-MS assessment will further undergo biopsy to rule out the presence of csPCa.

In this study, 19-BM again performed better than PSA and successfully classified most of PCa cases (19-BM: 42/48 patients; PSA: 32/48 patients). Moreover, both the NPV of 19-BM and the PPV were also higher than the values reported based on PSA performance The same observations were present when comparing to PSA density (AUC: 0.64) and the ERSPC3/4 calculator (AUC:0.67).

To investigate if improved performance can be achieved upon combination of 19-BM with current state-of-the-art risk calculators, several integrative diagnostics strategies were employed to develop DN including different combinations of 19-BM with the significant clinical variables like PSA, PSAD, age, ERSPC. In all above comparisons, the integrative DNs demonstrated improved performance, an observation which is in line with previous evidence for a level of complementarity of the diagnostics assays. Yet, in all comparisons the performance of the multimodal DNs did not significantly improve the 19-BM alone. An additional decision curve analysis was performed to assess the clinical benefit of the integrative model in comparison with the 19-BM, ERSPC 3/ 4, PSAD and PSA, demonstrating an improved net benefit particularly in the low range of risk threshold.

Considering the above scientific evidence as well as the very high NPV (>90%), the specific clinical impact of a non-invasive test like 19-BM or a DN based on 19-BM, would primarily be to guide and eventually reduce the number of invasive biopsies. The required high sensitivity for accurate detection of csPCa was achieved in this study. Upon potential application of such a test and in view of a positive test, urologists are alerted to perform a more thorough investigation, improving the overall accuracy in detection of csPCa. Lower specificity would likely result in more misclassifications of an insPCa form as a csPCa. As a result, a positive result based on 19-BM or a DN-based on 19-BM should be complemented by a biopsy procedure to rule out csPCa.

Considering the literature, several biomarkers have been tested to discriminate significant PCa, such as 4K score test, PHI, PCA3, SelectMDx [32, 33]. The PCA3 urinary assay based on post DRE samples demonstrated 67% sensitivity and 83% specificity for detecting PCa [34]. In comparison to the PHI in guiding initial and repeated biopsy, the PCA3 assay performed slightly, but not significantly inferior in both the initial (AUC_PCA3_: 0.57; AUC_PHI_: 0.69) and the repeated biopsy setting (AUC_PCA3_: 0.63; AUC_PHI_: 0.72) [35]. SelectMDx assay based on the combination of homeobox protein (DLX-1) and homeobox protein Hox-C6 (HOXC6), demonstrated an AUC of 0.73 [36], while Mi-Prostate Score which is based on the detection of the gene fusion TMPRSS2-ERG, in combination with urinary PCA3 resulted in an AUC of 0.76 for detection of PCa [37]. The validation results shown in this study with an AUC higher of 0.80 and 0.88 for the integrative nomogram, is higher than the range 0.57-0.73 which is shown by other biomarkers and justifies implementation of this approach, also in future investigative settings.

Although in this study previous potentially clinically useful positive results for 19-BM were replicated, the study also presents with certain limitations. Firstly, a direct comparison with the above biomarkers reported in the literature was unfortunately not possible, as paired data were not available. Moreover, this study was performed retrospectively, however, on samples that were prospectively collected. Based on the data presented, implementation of this approach, also in an investigative setting, seems to be highly justified. Additionally, and as another limitation, multiparametric MRI data was not available for this patient cohort. In line with the above, and in order to facilitate comparisons and inclusion of multiparametric MRI, assessment of complementarity with mpMRI in a future prospective setting is planned. However, based on the data already available, the potential use of this test in guiding PCa management should be considered as a valid option.

Collectively, the data presented in this study demonstrate the utility of a multimodal approach for improved non-invasive detection of significant PCa. Effective discrimination between clinically significant and insignificant PCa is expected to reduce the number of diagnostic biopsies and enable more frequent assessment of PCa (due to the non-invasive nature of the test), thus have a positive impact on PCa patient management, by improving patient compliance and reducing over-treatment and the associated costs. Considering the high NPV, the clinical utility of the presented nomogram could also be investigated in the context of guiding mpMRI. Along these lines, another study is planned to investigate the added value of this diagnostic nomogram to mpMRI in the context of detecting significant PCa.

## Data Availability

All data produced in the present study are available upon reasonable request to the authors

## Abbreviations List

AS: Active Surveillance
AUC: Area Under ROC Curve
CE: Capillary Electrophoresis
CI: Confidence Intervals
cs: clinically significant
DRE: Digital Rectal Examination
csPCa: clinically significant PCa
ERSPC: The European Randomized Study of Screening for Prostate Cancer
GS: Gleason score
MS: Mass Spectrometry
insPCa: insignificant PCa
PCa: Prostate Cancer
PSA: Prostate Specific Antigen
PSAD: Prostate Specific Antigen Density
ROC: Receiver Operating Characteristics
SVM: Support Vector Machine.

## ADITIONAL INFORMATION

### Ethics approval and consent to participate

This study was performed as part of BioGuidePCa project. Ethical approval was obtained by the Ethics Committee of Medical University Innsbruck (Innsbruck, Austria) in accordance with the Declaration of Helsinki and informed consent was obtained from all participants for the project.

### Funding

This work was supported by the BioGuidePCa (E! 11023, Eurostars) funded by BMBF (Germany).

